# Carotid Plaque Calcification Attenuation Characteristics are Associated with Intraplaque Hemorrhage Volumes: A 3D Segmentation-Based Analysis

**DOI:** 10.1101/2025.04.08.25325406

**Authors:** Jeremy Rubin, Quy Cao, Yu Sakai, Nathan Arnett, Huy Q. Phi, Andrew C. Hu, Brett L. Cucchiara, Daniel Bos, Luca Saba, Jarcy Zee, Jae W. Song

## Abstract

**Background and Purpose:** Despite the high prevalence of plaque calcifications in carotid atherosclerosis, the association between morphologic and attenuation features of calcifications and intraplaque hemorrhage (IPH) remains unclear.

**Methods:** Carotid bifurcation calcific plaques were identified on neck CTAs from patients with unilateral anterior circulation ischemic stroke consistent with embolic stroke of undetermined source. Plaque calcifications were manually segmented using 3D Slicer to measure volume, surface area, shape, and attenuation (Hounsfield Units (HU)) characteristics. IPH volume (IPHvol) was quantified using a semi-automated software. A linear mixed regression model evaluated associations between calcification features and IPHvol, adjusting for sex, age, and cardiovascular risk factors. An interaction term between calcification volume and attenuation was included after dichotomizing attenuation (>924HU) and volume (>30mm^3^) as high versus low based on median values.

**Results:** From 70 patients (median age 68 years, 50% female), 116 calcific plaques containing 269 plaque calcifications were analyzed. Adjusting for age, cardiovascular risk factors and plaque calcification features, being female showed lower IPHvols compared to males (mean ratio 0.34, p=0.002). A significant interaction between calcification volume and attenuation emerged (p=0.042). Among plaques with low volumes (<30mm^3^) of plaque calcifications, plaques with low-attenuation (<924HU) calcifications showed 5.53 times higher IPHvols than plaques with high-attenuation calcifications (p=0.003). Among plaques with high-attenuation calcifications (>924HU), plaques with high volumes of these calcifications showed 4.40 times higher IPHvols compared to low-volumes of high-attenuation calcifications (p=0.011).

**Conclusions:** Plaque calcification attenuation characteristics are associated with IPHvols. Beyond presence or volumes of plaque calcifications, calcification attenuation characteristics should be considered when evaluating unstable plaque components.

## Introduction

Atherosclerotic plaque calcifications are thought to be a hallmark of advanced vascular disease and aging. However, there is increasing evidence suggesting in addition to structurally stabilizing atherosclerotic plaque, plaque calcifications have a dual role and may also promote inflammation, associated with intraplaque hemorrhage (IPH) and specific patterns of plaque calcifications may be a marker for unstable plaque [1, 2]. In fact, differences in size, spatial distribution, and calcification-type may need to be considered [1]. For example, “spotty” calcifications may represent inflammatory calcific deposits that lead to rupture[3, 4], and have been shown to be predictive of cardiovascular events. Plaques with small low attenuating mineralizing calcific deposits may develop into niduses of microcalcifications, which have been linked to a higher risk of coronary heart disease [5]. These plaque types with spotty or microcalcifications may reflect higher risk plaque phenotypes. As calcifications coalesce and ossify into larger macrocalcifications, they may reflect a stabilizing fibrocalcific plaque phenotype [6]. Indeed coronary artery calcium density, weighted by CT attenuation characteristics, appears to be inversely associated with coronary heart and cardiovascular risk [7]. Plaque calcification features including attenuation characteristics, shapes, and sizes may thus shed insight into IPH, a marker of plaque instability [8].

We investigated the association of plaque calcification features with IPH volumes (IPHvol), in patients with embolic stroke of undetermined source (ESUS). Among patients with ESUS, mild to moderately stenotic noncalcified plaque have a prevalence of up to 25.8% of IPH in carotid arteries ipsilateral to the side of stroke [9]. Thus, patients with ESUS represent an ideal cohort to study the biologic interplay between plaque calcifications and IPH.

We leveraged 3D-volumetric imaging data and quantitatively analyzed each discrete plaque calcification on CTA neck exams from patients with ESUS. We hypothesized that carotid plaques with higher volumes (e.g., macrocalcifications) of low-attenuation, mineralizing plaque calcifications would be associated with higher IPHvols. Characterization of the relationships between these plaque components may shed insights into optimizing therapeutic strategies for high-risk atherosclerotic lesions.

## Methods

### Study Sample

The data that support the findings of this study are available from the corresponding author upon reasonable request. This retrospective study was approved by the local institutional review board. A retrospective observational cross-sectional study was conducted identifying consecutive patients with acute ischemic stroke admitted to our health system between October 1, 2015 – April 1, 2017 [10]. Patients were included if they were ≥18 years old, presented with unilateral anterior circulation ischemic stroke due to ESUS as determined by a vascular neurologist and imaged with CTA. Exclusion criteria included history of prior carotid endarterectomy or stenting, occlusion of either cervical internal carotid artery, absence of plaque calcifications or if the CTA neck was not within 10 days from the time the patient was last known normal.

### Image Acquisition and analysis

CTA was acquired in the axial plane using a 4th generation, helical CT scanner, slice thickness 0.75 to 1.5 mm, per institutional protocol. Iodinated contrast (100mL Isovue-370) was administered intravenously through a 20-gauge right antecubital catheter. All carotid plaque image analyses were performed across a 4cm segment across the carotid bifurcation. Each carotid plaque was processed using a semi-automated plaque composition segmentation software (Elucid Bioimaging, Boston, MA) to quantify intraplaque hemorrhage (IPH) volumes [11], as previously described [11, 12]. 3D Slicer (https://www.slicer.org) was used to manually label each discrete plaque calcification and derive calcification metrics. Two researchers were trained on 3D Slicer by a senior neuroradiologist using 5 practice cases. Following training, both researchers independently labeled each plaque calcification across a 4cm segment of the carotid bifurcation. Labels from each researcher were overlaid and proofed by a third independent radiologist. Quantitative values for each plaque calcification feature included: diameter (mm), volume (mm^3^), surface area (mm^2^), shape features (e.g., roundness, flatness, elongation) and attenuation values (Hounsfield units, HU). The number of discrete calcifications per plaque was also counted.

### Statistical Analysis

Normality was tested using the Shapiro-Wilk test. Summary statistics are reported as frequencies for categorical variables, medians and interquartile ranges for non-normally distributed variables and means and standard deviations for normally distributed continuous variables. Analyses were performed at the plaque-level and each carotid plaque within subject was treated as a separate observation as the goal was to investigate the biological association between each plaque calcification feature and the total IPHvol, which was the primary outcome. To derive a plaque-level summary statistic for calcification features, the mean of each calcification feature (e.g., volume, surface area, attenuation) within that plaque was calculated.

Linear mixed regression models assessed associations between plaque-level calcification features and IPHvol with a random intercept term for each subject to account for multiple plaques (right and left) within each subject. Analyses were adjusted for sex, age (above or below 60 years), and history of hypertension, diabetes mellitus, smoking and dyslipidemia. Multicollinearity was assessed using Pearson’s correlation coefficients between each pair of features. In addition, each feature’s variance inflation factor (VIF) was calculated and features with VIF>10 were sequentially removed from a linear regression model with IPHvol as the outcome. The model with lowest Akaike’s Information Criterion was used to determine the order in which features were removed. This process was repeated until all VIF values were below ten. The final set of features was used in the linear mixed regression models for IPHvol. An interaction term between calcification volume and attenuation was included after dichotomizing attenuation and volume as high versus low based on the median values. Specifically, calcification volume was considered high (e.g, macrocalcification) if above the median of 30mm^3^. Plaque calcification attenuation was considered high (e.g., ossifying calcification) if above the median of 924HU (**Figure 1**). Histograms of random intercepts and residual QQ plots were used to assess model assumptions. IPHvol showed residuals that were not normally distributed and IPHvols were thus natural log-transformed. Resulting regression coefficients were exponentiated and interpreted as mean ratios. Mean ratios represent a relative difference such that a value of 1 indicates no association with the outcome (IPHvol) and a value above 1 represents a positive association with the outcome. All statistical analyses were performed using R version 4.4.0.

**Figure 1:**
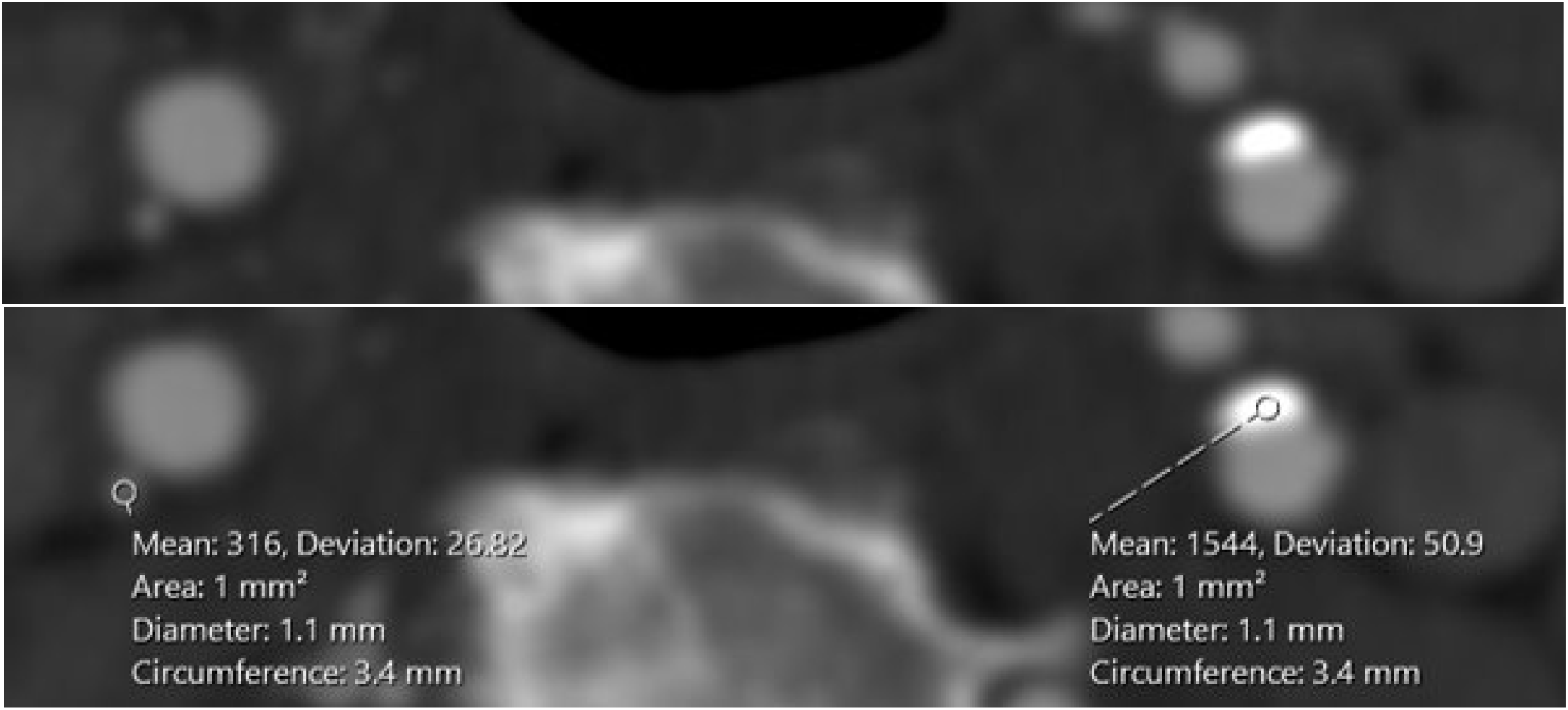
Examples of Plaque Calcifications with Low/mineralizing versus High/ossifying attenuation. As an example, the right carotid artery has a faint plaque calcification that measures a mean of 316 Hounsfield Units (HU) and is categorized as a plaque calcification with low attenuation (e.g., mineralizing). The left carotid artery plaque calcification has a plaque calcification that measures 1544HU and is considered high attenuation (e.g., ossifying).

## Results

From 70 subjects (mean age 68 (SD 11) years, 50% female), 116 calcific carotid artery plaques met inclusion criteria (**Figure 2**). **Table 1** reports the prevalences of hypertension, diabetes, dyslipidemia and smoking history in the cohort. **Table 2** summarizes plaque-level calcification features. Each carotid plaque had a median of 2 (IQR, 1-3) discrete calcifications.

**Table 1:** Demographics and Cardiovascular Risk Factors of Study Participants.

| **Characteristics (N=70 subjects)** |  |
| --- | --- |
| Age (years) | 68 (SD 11) |
| Female Sex | 35 (50%) |
| Hypertension | 53 (75%) |
| Diabetes Mellitus | 22 (31%) |
| Dyslipidemia | 28 (40%) |
| Smoking History | 14 (20%) |

Values are reported as mean (SD, standard deviation) or frequency (%).

**Table 2:**
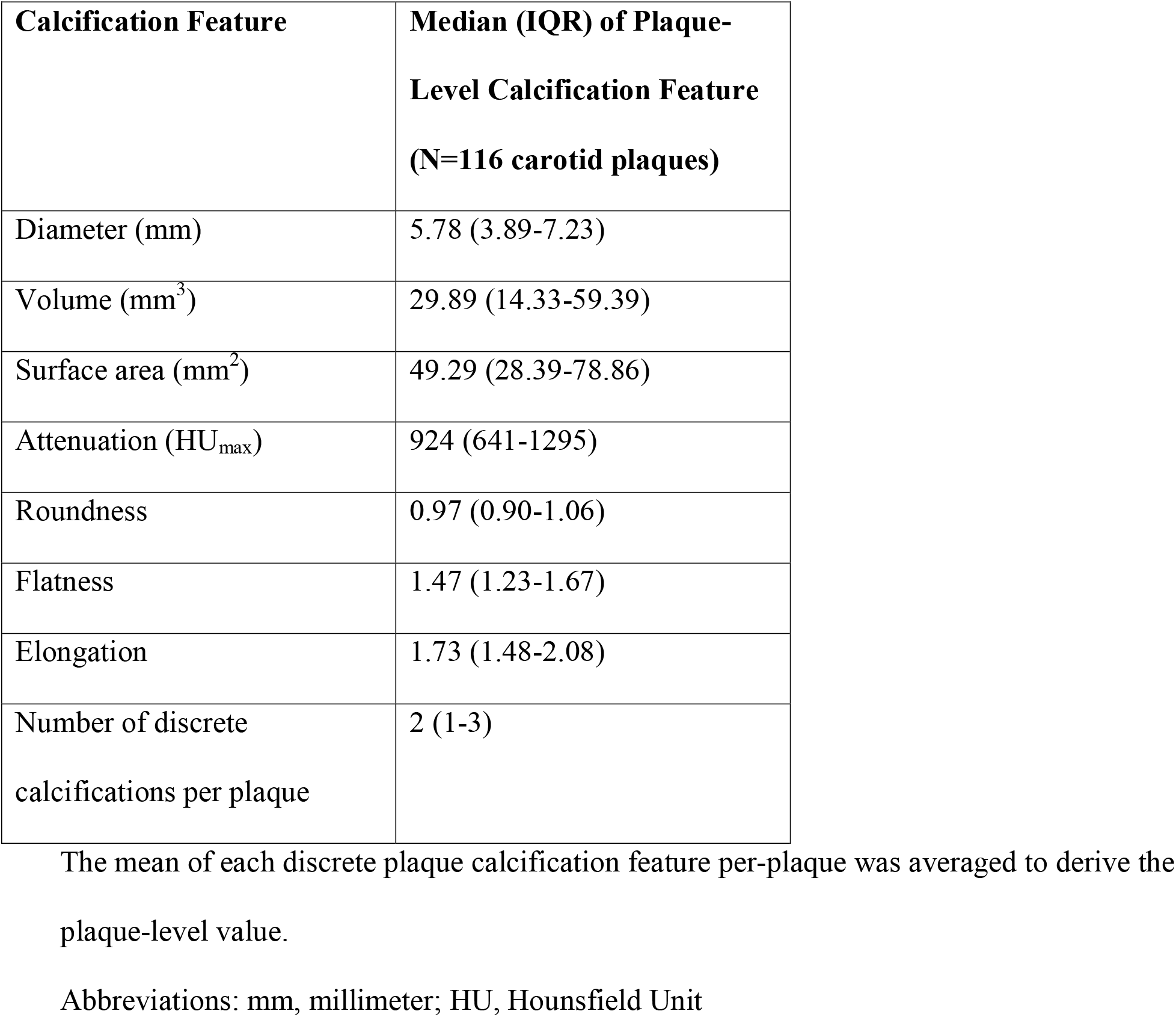
Summary of Calcification Features at the Plaque-Level.

| **Calcification Feature** | **Median (IQR) of Plaque-Level Calcification Feature**<br>**(N=116 carotid plaques)** |
| --- | --- |
| Diameter (mm) | 5.78 (3.89-7.23) |
| Volume (mm<sup>3</sup>) | 29.89 (14.33-59.39) |
| Surface area (mm<sup>2</sup>) | 49.29 (28.39-78.86) |
| Attenuation (HU<sub>max</sub>) | 924 (641-1295) |
| Roundness | 0.97 (0.90-1.06) |
| Flatness | 1.47 (1.23-1.67) |
| Elongation | 1.73 (1.48-2.08) |
| Number of discrete calcifications per plaque | 2 (1-3) |

The mean of each discrete plaque calcification feature per-plaque was averaged to derive the plaque-level value.
Abbreviations: mm, millimeter; HU, Hounsfield Unit

**Figure 2:**
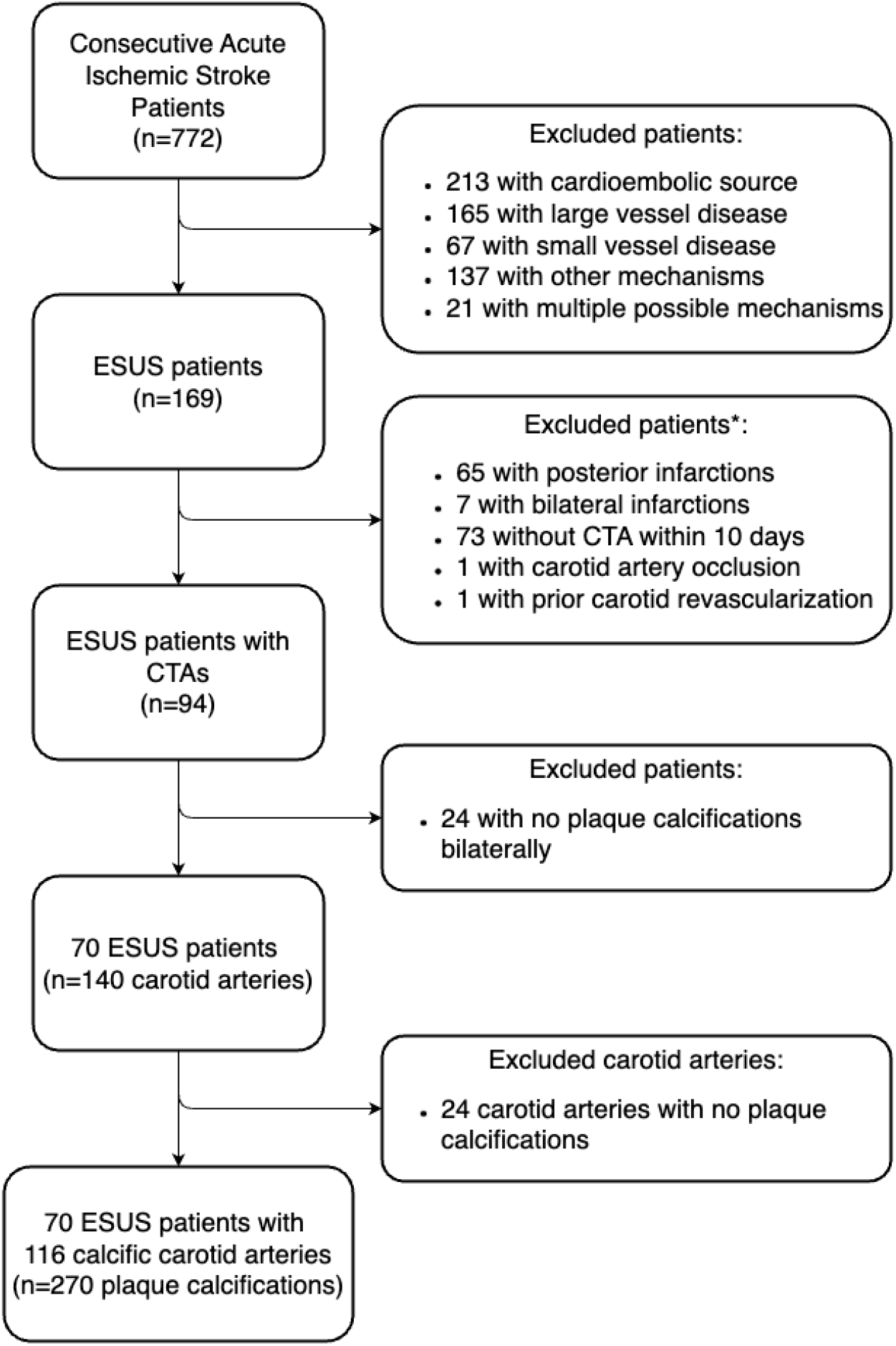
Flow of Patients with Calcified Carotid Plaque. From 772 patients with acute ischemic stroke, 70 patients met inclusion criteria. ^*^Reasons for exclusion are not mutually exclusive. Abbreviation: ESUS, embolic stroke of undetermined source

The regression results testing the associations between IPHvol with patient and plaque-level calcification features are shown in **Table 3**. Sex was significantly associated with IPHvol. Adjusting for age over 60, cardiovascular risk factors and other plaque calcification features, females had lower IPHvols than males (mean ratio 0.34, p=0.002).

**Table 3:**
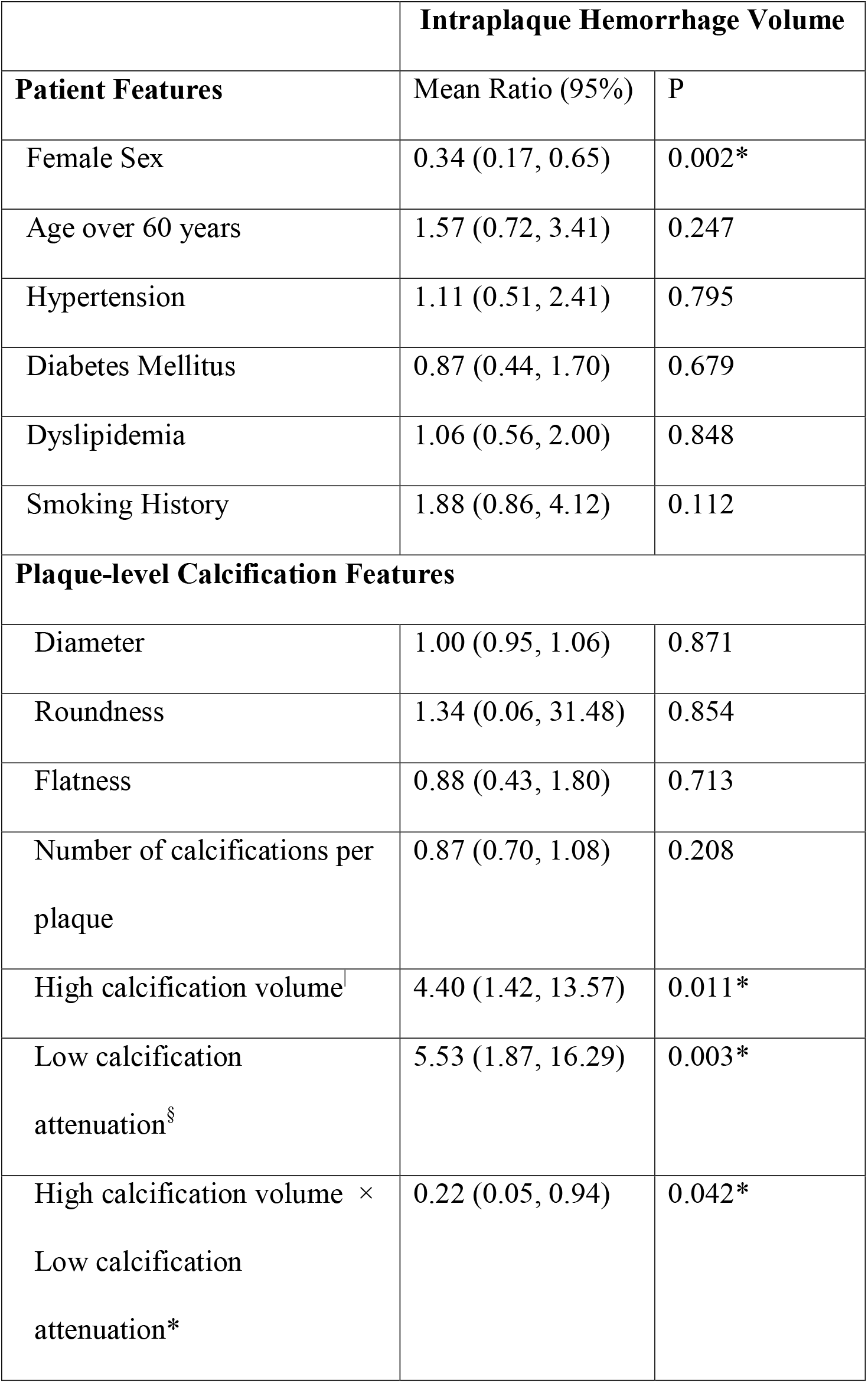
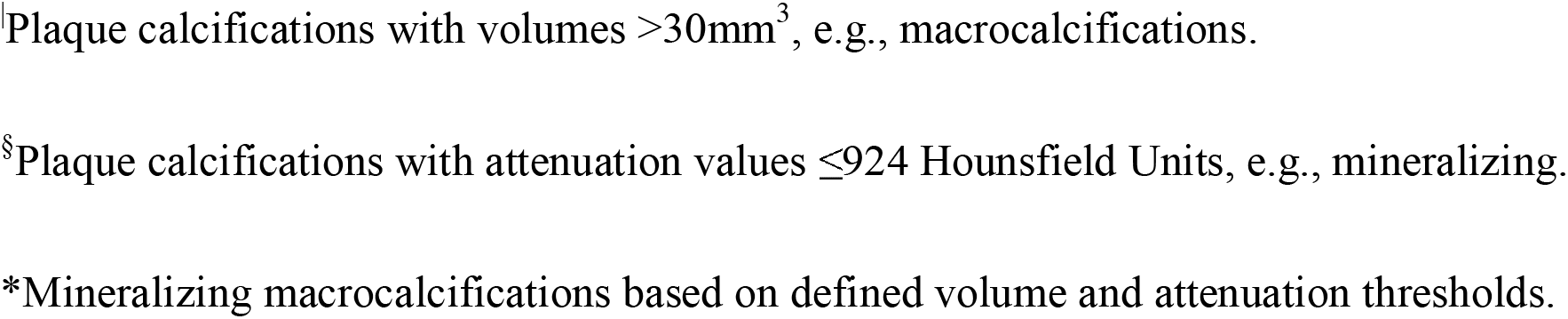
Associations between plaque-level calcification features and intraplaque hemorrhage volumes.

|  | **Intraplaque Hemorrhage Volume** |  |
| --- | --- | --- |
| **Patient Features** | Mean Ratio (95%) | P |
| Female Sex | 0.34 (0.17, 0.65) | 0.002* |
| Age over 60 years | 1.57 (0.72, 3.41) | 0.247 |
| Hypertension | 1.11 (0.51, 2.41) | 0.795 |
| Diabetes Mellitus | 0.87 (0.44, 1.70) | 0.679 |
| Dyslipidemia | 1.06 (0.56, 2.00) | 0.848 |
| Smoking History | 1.88 (0.86, 4.12) | 0.112 |
| **Plaque-level Calcification Features** |  |  |
| Diameter | 1.00 (0.95, 1.06) | 0.871 |
| Roundness | 1.34 (0.06, 31.48) | 0.854 |
| Flatness | 0.88 (0.43, 1.80) | 0.713 |
| Number of calcifications per plaque | 0.87 (0.70, 1.08) | 0.208 |
| High calcification volume<sup>‡</sup> | 4.40 (1.42, 13.57) | 0.011* |
| Low calcification attenuation<sup>§</sup> | 5.53 (1.87, 16.29) | 0.003* |
| High calcification volume × Low calcification attenuation* | 0.22 (0.05, 0.94) | 0.042* |

<sup>‡</sup>Plaque calcifications with volumes >30mm<sup>3</sup>, e.g., macrocalcifications.
<sup>§</sup>Plaque calcifications with attenuation values ≤924 Hounsfield Units, e.g., mineralizing.
\*Mineralizing macrocalcifications based on defined volume and attenuation thresholds.

An interaction between plaque calcification attenuation and volume was significant (p=0.042) accounting for sex, age over 60 years, cardiovascular risk factors, and other calcification features. Plaques with a low volume of low attenuation calcifications (e.g., mineralizing microcalcifications) showed 5.53 times higher IPHvols than plaques with a low volume of high attenuation calcifications (e.g., ossifying microcalcifications) (p=0.003). Plaques with high volumes of high attenuation calcifications (e.g., ossifying macrocalcifications) showed 4.40 times higher IPHvols than plaques with low volumes of high attenuation calcifications (e.g., ossifying microcalcifications) (p=0.011). In contrast, there was no significant association between attenuation and IPHvol among plaques with high calcification volume (mean ratio 1.23, p=0.698) nor between calcification volume and IPHvol among plaques with low attenuation calcifications (mean ratio 0.98, p=0.967).

IPHvols for plaques with low versus high plaque calcification attenuations and volumes were calculated using the average for continuous variables or most frequent categories for categorical variables for all other covariates in the model for IPHvol (**Figure 3**). Plaques with low calcification attenuation (e.g., mineralizing) had the highest IPH vols regardless of calcification volumes after controlling for sex, age, cardiovascular risk factors, and other calcification features.

**Figure 3:**
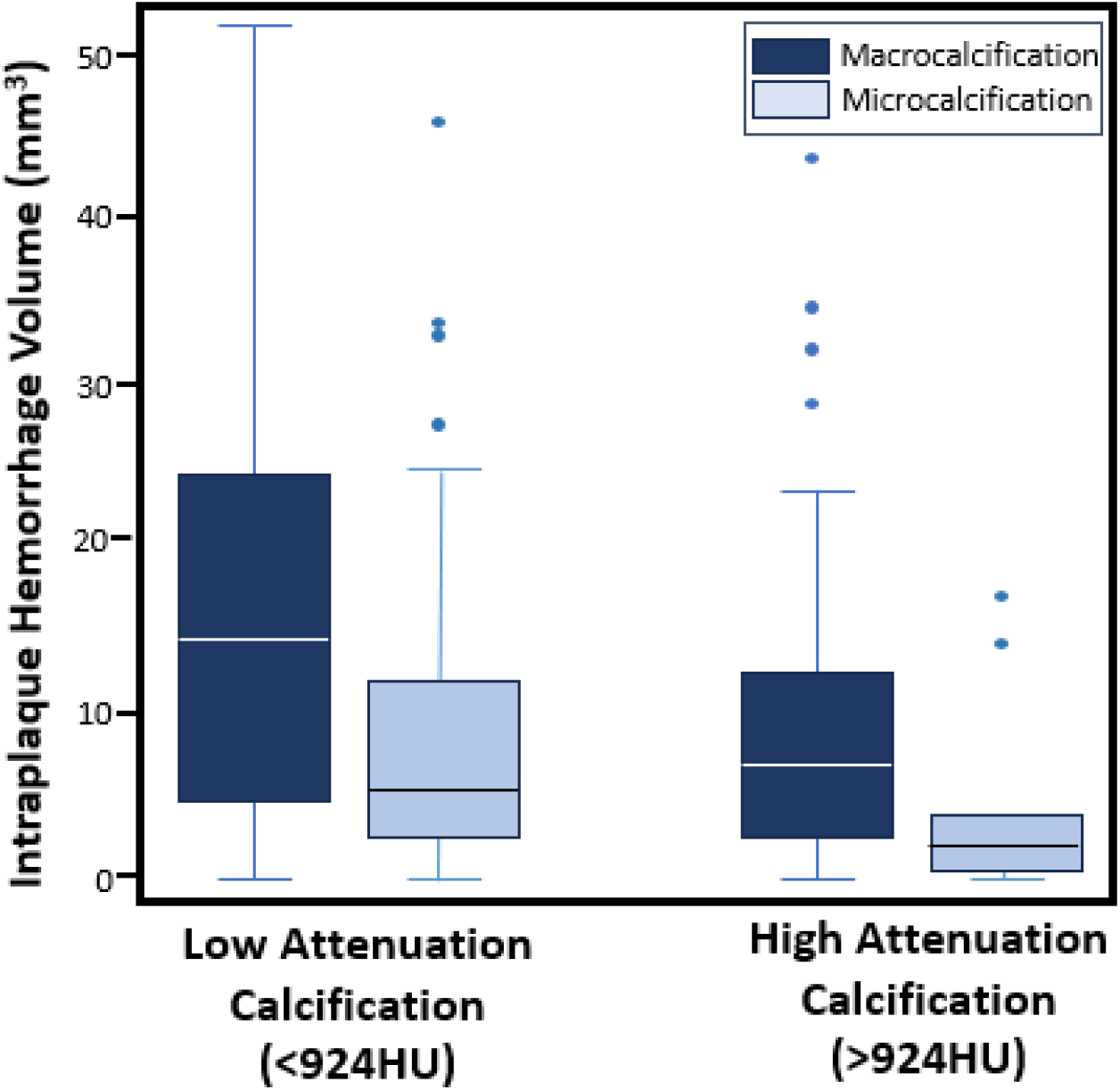
Intraplaque Hemorrhage Volumes by Subgroups of low/high Plaque Calcification Volumes and Attenuation Values. Subgroups based on plaque calcification attenuation (e.g., low HU/mineralizing vs high HU/ossifying) and volumes (e.g., macro versus microcalcification) and the intraplaque hemorrhage volumes are shown. Abbreviation: HU, Hounsfield Units

## Discussion

In our cohort of ESUS patients with calcific carotid plaques, we investigated plaque calcification features and their associations with IPHvol, a marker for plaque instability. Our results show that females had significantly lower IPHvols than males accounting for age, cardiovascular risk factors and plaque calcification features. Additionally, a significant interaction between plaque calcification attenuation and volume emerged in association with IPHvols. Lower attenuating plaque calcifications were associated with greater IPHvols. These results highlight the need to phenotype plaque calcification features, rather than simply noting the presence or volume of calcifications, as a potential imaging biomarker for plaque stability. Evidence increasingly suggests calcification in atherogenesis is a highly regulated process. Identifying the calcification features recognizable by noninvasive CT imaging may help recognize a substrate for treatment to stabilize or regress plaques [1].

Our results show a significant interaction between plaque calcification volume and attenuation. This raises the possibility of at least two distinct pathways contributing to plaque inflammation and IPH [13, 14]. Atherosclerosis initially develops as a fatty streak. As the lipid and necrotic debris from the fatty streak in early atherosclerosis accumulate, some components of the debris may mineralize into low attenuating and heterogeneous calcium deposits and microcalcifications. This pro-inflammatory milieu may drive the development neoangiogenesis and IPH formation. Our results showed that among plaques with low volumes of calcific plaque, low attenuation calcifications (e.g., mineralizing microcalcifications) had significantly higher IPHvols than high attenuation, ossifying calcifications. This result suggests an association between the low attenuation feature of plaque calcifications and an inflammatory microenvironment. Subsequently, while in this proinflammatory milieu, the low attenuation calcium deposits may coalesce into larger plaque calcifications. In fact, macrophages are reported to control the mineralization process of plaque calcifications from microcalcifications to osteoid metaplasia of bone-like tissue [15], which is likely to show higher CT-attenuation characteristics. The larger sized and highly attenuating calcifications (e.g., ossifying macrocalcifications) may additionally promote mechanical stress on the overlying endothelium and fibrous cap, leading to micro-disruptions, fissures, vascular leakage and IPH accumulation. Our results also show that among plaques with highly attenuating ossifying plaque calcifications, larger calcific volumes showed significantly higher IPHvols than plaques with smaller volumes. One explanation for this result is that larger highly attenuating plaque calcifications (e.g., ossifying macrocalcifications) may induce mechanical stress if adjacent to the endothelium and cause plaque rupture.

Our study also indicates that sex as a biological variable has a strong influence in plaque composition. Many lines of work have shown that women tend to have a lower prevalence of inflammatory plaque components compared to men of the same age [12, 16]. A comparison of 156 men and 68 women from the PARISK (Plaque at Risk) study, which included patients with recent ischemic cerebrovascular symptoms and <70% ipsilateral carotid stenosis, showed women have significantly lower total plaque volumes and men were more likely to have IPH than women, adjusting for total plaque volumes (OR=2.8 [95%CI, 1.3-6.3]). Plaque in men also showed higher complexity with the coexistence of calcifications and lipid-rich necrotic cores and IPH compared to women (OR=2.7 [95%CI, 1.1-5.9]) [17]. Studies should consider sex when interpreting plaque composition results and efforts should be made to recruit balanced cohorts of men and women.

There are several limitations to this study. First, each plaque calcification was manually segmented and assessed visually, which could lead to errors in segmentation. To minimize errors, 2 raters independently segmented each carotid plaque, and a third radiologist used both segmentation masks to proof and produce a final high-quality segmentation dataset. Second, we used conventional energy integrating detector CT technology, a soft tissue kernel and lower spatial resolutions (0.75-1.5mm), which is our institutional protocol for CTA neck vascular imaging. Calcifications may bloom on the soft tissue kernel, which may lead to oversegmentation of calcification volumes. Additionally, blooming artifact may lead to a cluster of discrete calcifications appearing contiguous rather than non-contiguous. Future investigations will explore differences in calcification features using a sharp bone kernel on ultra-high spatial resolution images (0.2mm), which is available with photon-counting CT [18]. Third, a CT-based semi-automated segmentation software identified and measured IPHvols, as used in other studies [11, 12, 19]. No carotid vessel wall MR or histologic specimens was available for IPH validation in this. A future direction is to investigate photon □ counting spectral CT technology to distinguish plaque components. Fourth, high versus low attenuation calcification groups were based on median HU values of this patient cohort. Low versus high attenuation HU ranges could reflect different elemental calcium, such as calcium hydroxyapatite versus calcium oxalate, which have been associated with unstable versus stable carotid plaques, respectively [20, 21]. Further validation of these HU ranges for different calcium types using spectral CT would be informative. Finally, our study was cross-sectional and conclusions based on correlation rather than causation.

In conclusion, we show a significant interaction between plaque calcification attenuation and volumes and their association with IPHvols. Understanding the biologic interplay and the role of low attenuating plaque calcifications may provide insight into potential therapeutic targets to stabilize or regress high-risk inflammatory plaque phenotypes.

## Data Availability

All data produced in the present study are available upon reasonable request to the authors.

## Acknowledgement and Disclosure

None.

